# SARS-CoV-2 Detection in the Nasopharyngeal Swabs and Saliva of College Students using RT-qPCR and RT-LAMP

**DOI:** 10.1101/2021.03.31.21254634

**Authors:** D. A. Bikos, C. Hwang, K. A. Brileya, A. Parker, E. K. Loveday, M. Rodriguez, I. Thornton, T. LeFevre, J. N. Wilking, M. Dills, S. T. Walk, A. K. Adams, R. K. Plowright, A. B. Hoegh, J. R. Carter, J. Morrow, M. P. Taylor, D. E. Keil, M. W. Fields, C. B. Chang

## Abstract

**Background:** Diagnostic testing can identify outbreaks and inform preventive strategies for slowing the spread of SARS-CoV-2, the virus that causes Covid-19. The “gold standard” method for detection of SARS-CoV-2 is reverse transcription quantitative polymerase chain reaction (RT-qPCR) performed on samples collected using nasopharyngeal (NP) swabs. While NP RT-qPCR achieves high sensitivity, it requires trained personnel to administer and suffers from lengthy time-to-result. Instead, rapid saliva-based reverse transcription loop-mediated amplification (RT-LAMP) screening methods may offer advantages in sample collection and speed.

**Methods:** Regardless of symptomatic presentation, a total of 233 individuals were tested for SARS-CoV-2 using NP RT-qPCR, alongside saliva-based RT-qPCR (SalivirDetect) and RT-LAMP (SLAMP), a simple and rapid fluorometric RT-LAMP assay performed directly on heat-inactivated saliva without any additional treatments or RNA extraction. SLAMP is conducted in triplicate and takes 45 min. Samples found negative using both saliva-based methods but positive under CDC NP RT-qPCR above the saliva method LoD were excluded from evaluation, suggesting significant differences in viral titer between sampling sites. Individuals who consumed potential inhibitors in the form of food, drink, and oral health products within 30 min of sampling were identified using a self-reported questionnaire.

**Results:** Of the 233 NP RT-qPCR tests, 58 were positive and 175 were negative. Comparatively, SLAMP resulted in 95% sensitivity and 98% specificity and SalivirDetect 97% sensitivity and 98% specificity. Prior consumption had no measurable effect on test outcomes, except for drinking, which lowered Ct values in saliva.

**Conclusions:** SLAMP requires less technician and instrument time than CDC-approved NP RT-qPCR and demonstrates that saliva-based RT-LAMP can enable frequent and rapid identification of pre-symptomatic and asymptomatic SARS-CoV-2 infections with high sensitivity and specificity.

## Introduction

The SARS-CoV-2 virus^1^ emerged in late 2019, rapidly developing into a worldwide pandemic still posing a persistent threat to public health, economics, and quality of life. While real-world vaccine efficacy continues to be evaluated^2^, uncertainties over variant escape^3^, vaccine availability^4^, and duration of immunity^5^ suggest that testing will continue to play an indispensable role in managing the disease into the future. Viral spread can be controlled by deploying diagnostic tests that rapidly identify infected individuals for quarantine and contact tracing. Rapid and frequent SARS-CoV-2 testing is needed to identify new outbreaks as the world struggles to lift lockdowns^6^ and reopen schools for in-person instruction in Fall 2021^7^.

The “gold standard” method for detecting SARS-CoV-2 has been reverse transcription quantitative polymerase chain reaction (RT-qPCR) performed on samples collected using nasopharyngeal (NP) swabs^8^. However, RT-qPCR testing requires significant reagent consumption, specialized equipment, trained operators, and several hours to perform. Collection of NP swab samples is invasive and must be performed by trained medical personnel. Performing surveillance at scale calls for innovative testing strategies that are inexpensive, minimize reagent consumption, decrease assay time-to-result, and avoid restrictions in available personnel^9,10^.

One such alternative to RT-qPCR is reverse transcriptase loop-mediated isothermal amplification (RT-LAMP), an isothermal technique for the amplification of RNA^11^. RT-LAMP simplifies SARS-CoV-2 testing by eliminating the long assay times and technical barriers such as the electronically controlled thermal cycling at high temperatures required by PCR-based methods^12-17^. RT-LAMP is substantially faster than RT-qPCR, and when performed on saliva samples^18^, eliminates the need for specialized swabs and operators while assuaging the public reluctance to testing, largely brought on by invasive NP swabs^19^. The RT-LAMP assay is usually completed within 45 min to confirm a negative, while some exceptionally high viral loads become detectable at ≈10 min^20^.

Here, we report the findings of a multi-day pilot study to evaluate saliva-based SARS-CoV-2 detection using RT-qPCR and RT-LAMP methods compared alongside to Centers for Disease Control and Prevention (CDC)-approved NP RT-qPCR testing at Montana State University (MSU) during mid-November 2020, a time when Covid-19 cases reached county-wide highs (Figure S1). We present the results of SLAMP, a rapid fluorometric RT-LAMP assay performed directly on saliva without chemical extraction steps. Additionally, we compare SLAMP to SalivirDetect^21^, a direct saliva-to-RT-qPCR assay and both saliva tests are compared to CDC-approved RT-qPCR performed on samples swabbed from the nasopharynx. Each participant was provided a self-reported survey identifying samples collected within 30 min of consuming food, drink, or oral hygiene products to determine the extent to which saliva test results might be affected.

## Methods

### Participants

Samples were collected from 233 participants who visited the on-campus CDC-approved RT-qPCR testing site provided for persons exhibiting Covid-19 symptoms or who suspected recent contact with SARS-CoV-2-positive individuals. We directly compared samples from two sources on the body of each participant, NP swabs, tested using CDC-approved RT-qPCR, and saliva, tested using SLAMP and SalivirDetect. All individuals tested signed a consent form before participating in this study (Supplementary Information – Consent Form).

### Self-Reported Questionnaire

Participants were provided with a self-reported questionnaire (Supplementary Information - Questionnaire) that collected demographic information including age, gender, race, and ethnicity. Additionally, subjects were asked to confirm whether they were currently symptomatic or asymptomatic and whether they had recent contact with infected individuals. Finally, subjects were asked to disclose if they had, in the last 30 min, consumed food or liquid, used mouthwash, gum or lozenges, smoked, vaporized or chewed tobacco, or brushed their teeth.

### Sample Collection

All sampling occurred at an outdoor parking lot in drive-through format while participants remained inside automobiles as an additional precaution against viral transmission. Individuals who expressed interest in participating in our pilot study were administered NP swabs by trained staff before donating an additional saliva sample as outlined by the university IRB (Supplementary Information – IRB).

### Viral Inactivation - NP Swab

NP swabs were heat-inactivated by diluting collection buffer 1:2 with molecular biology grade H_2_O (50 μL buffer: 50 μL H_2_O) and assigned a location on a 96-well PCR plate. Plates were covered with sealing foil and heated to 95 °C for 5 min in a standard thermocycler before being cooled to 4 °C until ready to test.

### Viral Inactivation - Saliva

In the case of saliva collection, participants were given a 3D-printed accessory caddy (Supplementary Methods, Figure S2) containing a 30-mL polypropylene medicine cup (MedPride 97205), a generic 1-mL transfer pipet, and a screw cap tube (VWR 16466-040). Saliva was expressed into the medicine cup (Figure S3i) before using the pipet to transfer ≈1.0 mL into the screw-cap tube (Figure S3ii). Tubes were indexed with heat- and water-resistant adhesive labels (Electronic Imaging Materials, Inc. 667) prepared via barcode printer (TSC MB340T). Samples were inactivated using a heat block (Labnet AccuBlock) set to 95 °C for 15 min (Figure S3iii) simultaneously during which time ribonucleases were denatured and virions lysed. While not strictly necessary, samples were removed from heat treatment and left at room temperature for 20 min to allow debris in the saliva to settle for easier pipetting. Barcodes were entered into records using a handheld scanner (Motorola Symbol LS2208-SR20007R-NA).

### CDC-approved RT-qPCR from NP Swab

Next, 10 uL of diluted, heat-inactivated^22^ sample material was mixed with 15 μL of each 1-step PCR master mix consisting of: 1.) 1.5 μL each N1 and N2 primer probe set, 5 μL Quantabio Ultraplex 1-Step Toughmix (4×), and 7 μL H_2_O per reaction; 2.) 1.5 μL human RNase P reaction primer probe mix, 5 μL Quantabio Ultraplex 1-Step Toughmix (4×), and 8.5 μL H_2_O per reaction. Real-time PCR thermocycling was performed per CDC guidelines^23^. Each sample was screened in a combined N1, N2 reaction with an internal control human RNase P reaction (Supplementary Methods). Samples with no detectable fluorescence for either assay and those with SARS-CoV-2 target fluorescence between 39.5 and 45 cycle thresholds (Ct) were re-tested using a validated RNA purification kit (Promega Maxwell RSC Viral Total Nucleic Acid Multi-Pack Kit, ASB1330). RNA-purified samples were assayed under the same conditions, but with 5 μL of purified sample added to each reaction for 20 μL total reaction volume. No replicates were performed.

### SalivirDetect - RT-qPCR in saliva

SLAMP was compared to the saliva-based RT-qPCR assay SalivirDetect with FDA Emergency Use Authorization (EUA) application number EUA202615, submitted August 25, 2020 and developed by Drs. Phillip Buckhaults and Carolyn Banister (University of South Carolina). SalivirDetect, like SLAMP, uses heating at 95 °C to process saliva without inactivation buffers or additives. SalivirDetect was conducted in the InHealth Life Sciences CLIA/CAP laboratory led by Dr. Deborah Keil (MSU). Briefly, ≈5 mL of saliva was collected in a 50-mL centrifuge tube. The tube was placed in a 95 °C oven (Fisherbrand Isotemp General Purpose Heating and Drying Oven) for 45 min, then allowed to sit at room temperature to cool. Two 5-μL aliquots of the saliva were transferred to two wells in a 96-well plate containing Luna Kit RT-qPCR reagents (E3600, New England Biolabs), one well containing N1 primers and the second well containing human RNase P primers from the United States CDC Real-Time Reverse Transcription PCR Panel for SARS-CoV-2 detection (Supplementary Methods). Samples were run on a CFX Opus 96 Real-Time qPCR instrument (Bio-Rad, cat no. 12011319). Only a single test per sample was performed with no replicates.

### SLAMP - RT-LAMP in saliva

RT-LAMP reactions were set up as described by protocol E1700 (New England Biolabs) at 25 μL final reaction volume with the following modified formulation: 12.5 μL WarmStart 2X Master Mix (E1700L, New England Biolabs) 0.175 μL of dUTP (N0459S, New England Biolabs), 0.5 μL of UDG (M0372L, New England Biolabs), 2.5 μL of duplex NE primer mix (Supplementary Methods), 0.5 μL of 25-μM SYTO-9 (S34854, Invitrogen), 0.25 μL of 25-μL ROX reference dye (61110, Lumiprobe), 2.5 μL of 400-mM molecular biology grade guanidine HCl (GuHCl) (J65661, Alfa Aesar), 1.075 μL of nuclease-free water (B1500L, New England Biolabs), and 5 μL of heat-inactivated saliva sample. Controls were run on each 96-well plate, including SARS-CoV-2-negative heat-inactivated saliva as a no-template control (NTC), human beta-actin (ACTB) as a positive internal control, and three positive test controls of 5 × 10^4^, 5 × 10^2^, and 5 × 10^0^ copies/μL synthetic SARS-CoV-2 RNA (NIST, RGTM 10169 Fragment 1) (Supplementary Methods). Positive test control dilutions were made using heat-inactivated SARS-CoV-2-negative saliva. All SLAMP reactions were prepared in 96-well qPCR plates (Applied Biosystems, 4483485) sealed using an ALPS 50V manual heat sealer (Thermo Scientific) with Clear Seal Diamond films (AB-0812, Thermo Scientific). Reactions were performed at 65 °C (1.6 °C/s ramp) on a QuantStudio 3 Real-Time PCR System (Applied Biosystems) (Figure S3iv) for 45 min. Fluorescence measurements for SYTO-9 and ROX channels were recorded every 1 min. SARS-CoV-2 may be present in highly variable amounts within the nasopharynx and the saliva^24^. To avoid invalid test samples where RNA was present only in the NP swab, only NP RT-qPCR-positive samples that also tested positive under at least one of the two saliva methods were included in this study. Any time-to-positive Tp > 40 min was considered negative. See Supplementary Methods for reaction curve-fitting procedure and SLAMP formulation optimization. Each saliva sample was run in triplicate.

### Statistical Analysis

Statistical bootstrapping^25^ methods can predict how the normally triplicate SLAMP sensitivity and specificity may change when performed only in duplicates or single tests. Using our (triplicate, 45 min) SLAMP test results, 1,000 random bootstrap samples were used to predict sensitivity with respect to CDC-approved NP RT-qPCR at 95% confidence intervals when only 1 or 2 SLAMP reactions are performed per sample. In addition, we model the outcomes of SLAMP testing under hypothetical disease prevalence scenarios. Based upon the experimental sensitivity of SLAMP in triplicate and the bootstrap-predicted sensitivities of duplicate and single SLAMP tests, we determine the total number of detectable positive individuals based on a daily testing capacity of six 384-well plates, reserving 10 wells for controls when performed in triplicate, duplicate, or single tests. Specificity is assumed to remain constant.

All t-tests are reported as p values. Pearson correlation coefficients are reported as r values.

## Results

### Comparing testing methods: SLAMP, SalivirDetect, and NP RT-qPCR

We compare samples from two matrices, NP swab and saliva, across three different testing methods, SLAMP, SalivirDetect, and the “gold standard” CDC-approved NP RT-qPCR, the latter performed using NP swab while saliva, shown to provide sufficient viral RNA for detection^26,27^, was used for both SLAMP and SalivirDetect. Both SalivirDetect and CDC-approved NP RT-qPCR are PCR-based while SLAMP uses isothermal LAMP amplification. SLAMP was performed in triplicate while SalivirDetect and NP RT-qPCR tests were performed only once (Figure 1).

**Figure 1.**
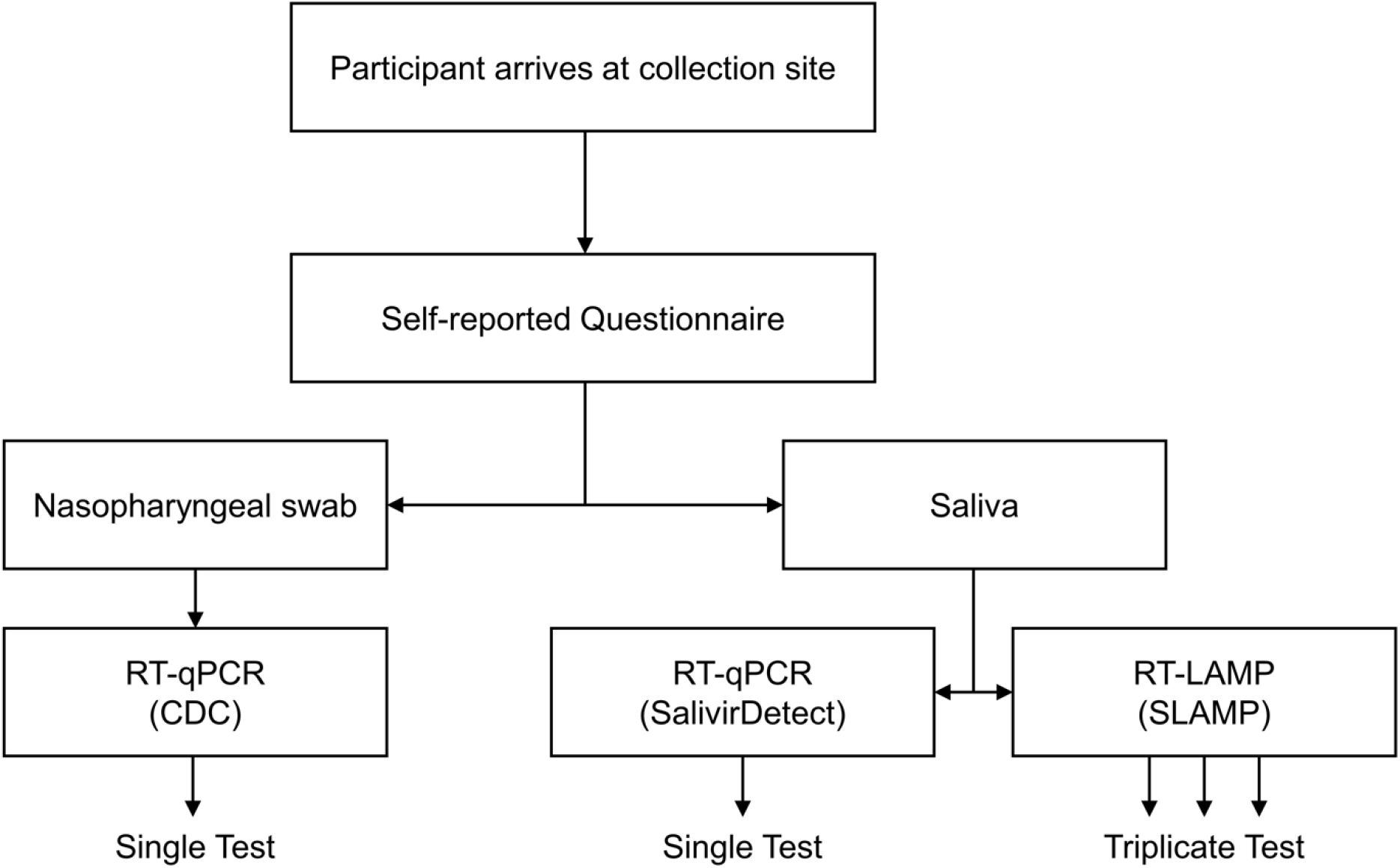
Sample collection and testing.

### Demographics

Participant ages ranged from 17 to 63 with a median age of 20. Males made up 47% and females 53%. Participants were comprised of 94% white and 6% other races including Asian and mixed-race individuals of American Indian, Alaskan Native, Native Hawaiian or Pacific Islander, and African American descent. Ethnically, 9 individuals self-reported as Hispanic or Latino, 207 as not Hispanic or Latino, and 17 chose not to respond (Table 1).

**Table 1.** Self-reported Demographic Information and Clinical Presentation.

|  |  |
| --- | --- |
| **Characteristic** | **Participants**<br>**(N = 233)** |
| **Sex - no.** |  |
| Male | 109 |
| Female | 124 |
| **Median Age (IQR)\* – yr** | 20 (19-22)<sup>†</sup> |
| **Race – no.** |  |
| White | 220 |
| Asian | 1 |
| Mixed race | 11 |
| Not provided | 1 |
| **Ethnicity – no.** |  |
| Hispanic or Latino | 9 |
| Not Hispanic or Latino | 207 |
| Not provided | 17 |
| **Clinical Presentation – no.** |  |
| Symptomatic (known exposure) | 76 |
| Symptomatic (no known exposure) | 88 |
| Asymptomatic (known exposure) | 64 |
| Asymptomatic (no known exposure) | 4 |
| Not provided | 1 |
| **Consumption within 30 min of Saliva Sampling<sup>††</sup> – no.** |  |
| Only Food | 4 |
| Only Drink | 23 |
| Both Food and Drink | 10 |
| Neither Food nor Drink | 21 |
| **Mouthwash<sup>†††</sup>** |  |
| Yes | 5 |
| No | 52 |
| **Gum or Lozenge<sup>†††</sup>** |  |
| Yes | 6 |
| No | 52 |
| **Tobacco<sup>†††</sup> (Smoke, Vape, or Chew)** |  |
| Yes | 7 |
| No | 51 |

| Brushed Teeth<sup>†††</sup> |  |
| --- | --- |
| Yes | 31 |
| No | 27 |

\* Interquartile range.
† N = 223 (10 non-respondent).
†† Tested positive using SalivirDetect.
††† Multiple selections possible.

### Clinical presentation

Participants self-reported their illness presentation as either symptomatic or asymptomatic and indicated whether they had knowledge of recent exposure to a confirmed Covid-19-positive individual. A majority 74% of study participants self-reported as symptomatic among whom 46% indicated knowledge of recent exposure. Among the 29% asymptomatic, 6% were unaware of exposure, the remaining 94% sought testing after learning of SARS-CoV-2 exposure (Table 1).

### Potential saliva contamination

Testing was conducted on a first come, first served basis without appointments. Therefore, instructions to abstain from food, drink, or oral hygiene products before donating saliva could not be provided to participants ahead of time. To determine the extent to which saliva may have been affected, the questionnaire asked participants whether they had eaten food or consumed drinks within 30 min of sampling. In addition, other consumables or oral hygiene products that may have persisted in saliva were also included, namely mouthwash, gum/lozenge, tobacco (whether smoked, vaporized, or chewed), and toothpaste from brushing teeth. Many participants had consumed multiple items. In order to make quantitative comparisons between saliva sample Ct values, we report the consumption survey results for individuals who testing positive using SalivirDetect (Table 1).

### Testing outcomes - performance of the three methods

Performance metrics of saliva methods SalivirDetect and SLAMP were evaluated by comparing results to the “gold-standard” CDC-approved NP RT-qPCR method between 233 individuals tested using all three methods. We report the true positive (TP), true negative (TN), false positive (FP), and false negative (FN) counts along with the true positive rate (TPR) and true negative rate (TNR), also called sensitivity and specificity, respectively (Table 2). The sensitivity/specificity (in %) of SalivirDetect and SLAMP methods were 97/98 and 95/98, respectively. Based on NP RT-qPCR, 58 positive and 175 negative individuals were identified resulting in a disease prevalence of 25%.

**Table 2.** Test Outcomes and Performance Metrics.

| Table 2. Test Outcomes and Performance Metrics. |  |
| --- | --- |
| Method and Category |  |
| NP RT-qPCR (CDC-approved "gold standard") |  |
| Diseased | 58 |
| Non-diseased | 175 |
| Saliva RT-qPCR (SalivirDetect) |  |
| True Positives (TP) | 56 |
| True Negatives (TN) | 220 |
| False Positives (FP) | 3 |
| False Negatives (FN) | 2 |
| Sensitivity (%) | 97 |
| Specificity (%) | 98 |
| Saliva RT-LAMP (SLAMP) |  |
| True Positives (TP) | 55 |
| True Negatives (TN) | 172 |
| False Positives (FP) | 3 |
| False Negatives (FN) | 3 |
| Sensitivity (%) | 95 |
| Specificity (%) | 98 |

### Comparing positive samples - two sample matrices and three testing methods

The 58 samples found positive under CDC-approved NP RT-qPCR are placed in order of increasing Ct value (decreasing effective SARS-CoV-2 genome concentration or viral titer) and compared to Ct values under SalivirDetect (Figure 2A) and Tp values using SLAMP (Figure 2B). Samples positive under saliva-based tests are compared in order of increasing SalivirDetect Ct value (Figure 2C) and increasing SLAMP Tp value (Figure 2D). False negatives are shown as missing bars and indicated by black arrows.

**Figure 2.**
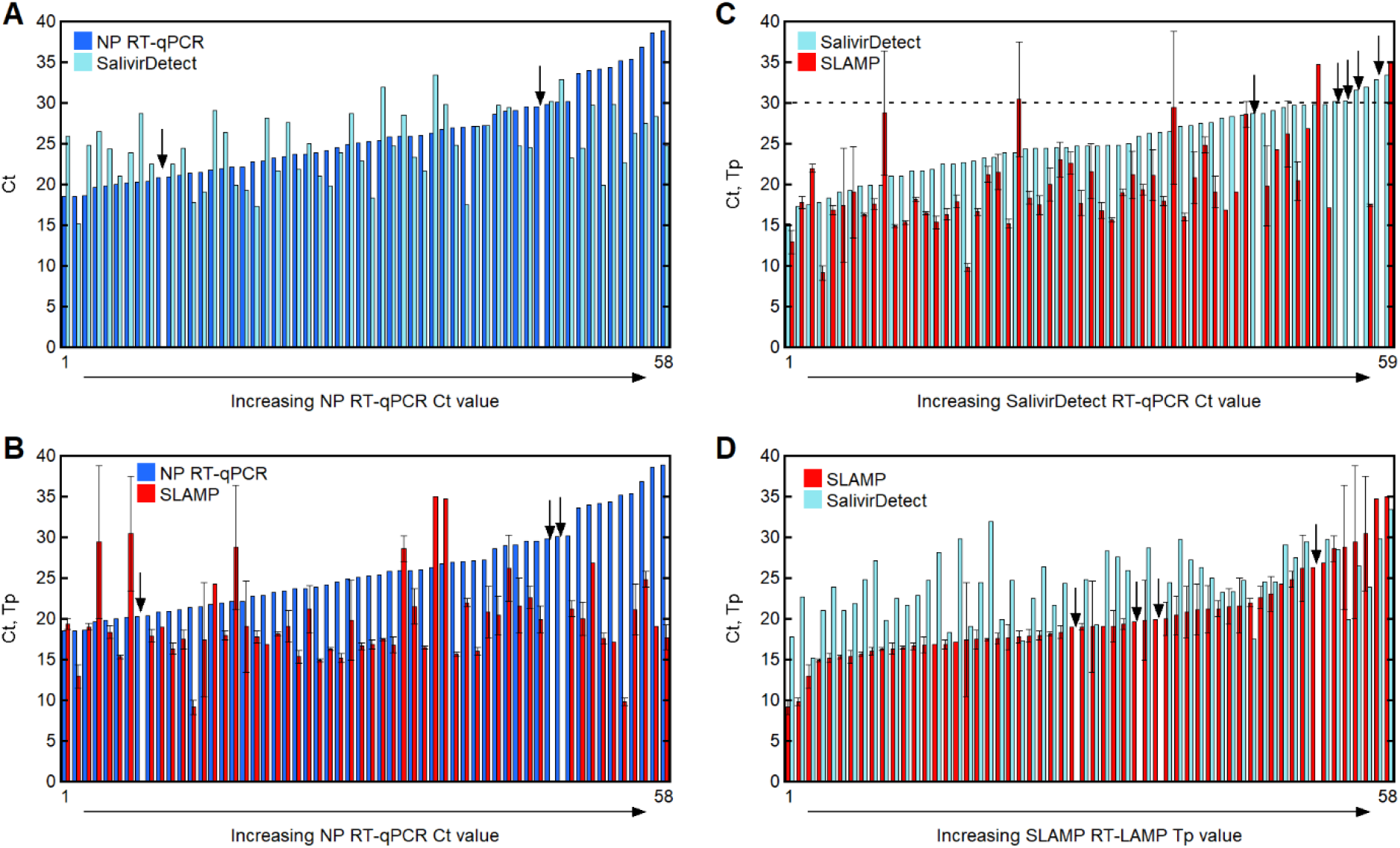
Positive sample results compared between three tests. Ct values of N=58 positive NP RT-qPCR and SalivirDetect samples in order of increasing CDC-approved NP RT-qPCR Ct value (Panel A). Ct values of N=58 positive CDC-approved NP RT-qPCR and Tp values of SLAMP samples in order of increasing CDC-approved NP RT-qPCR Ct value (Panel B). Ct values of N=59 positive SalivirDetect and Tp values of SLAMP samples presented in order of increasing SalivirDetect Ct value. Dashed line indicates 95% confidence interval SLAMP limit of detection (Panel C). Tp and Ct values of N=58 positive SLAMP and SalivirDetect tests, respectively, in order of increasing SLAMP Tp value (Panel D). Black arrows indicate undetected samples. Error bars represent one standard deviation.

### Test outcomes and viral titers by age

A total of 223 participants provided age information and among these 56 were confirmed positive for SARS-CoV-2 using CDC-approved NP RT-qPCR. A histogram of these ages shows the positive results (gray) superimposed over the negative results (white), including one outlier age of 63 (Figure 3A). The most common age to be positive and negative is equal to the mode of 19 years old. Not surprisingly, the positive and negative distributions are similar (p = 0.407). Average Ct values as a function of age also indicate no trend in positivity as a function of this narrow age window (Figure 3A, inset).

**Figure 3.**
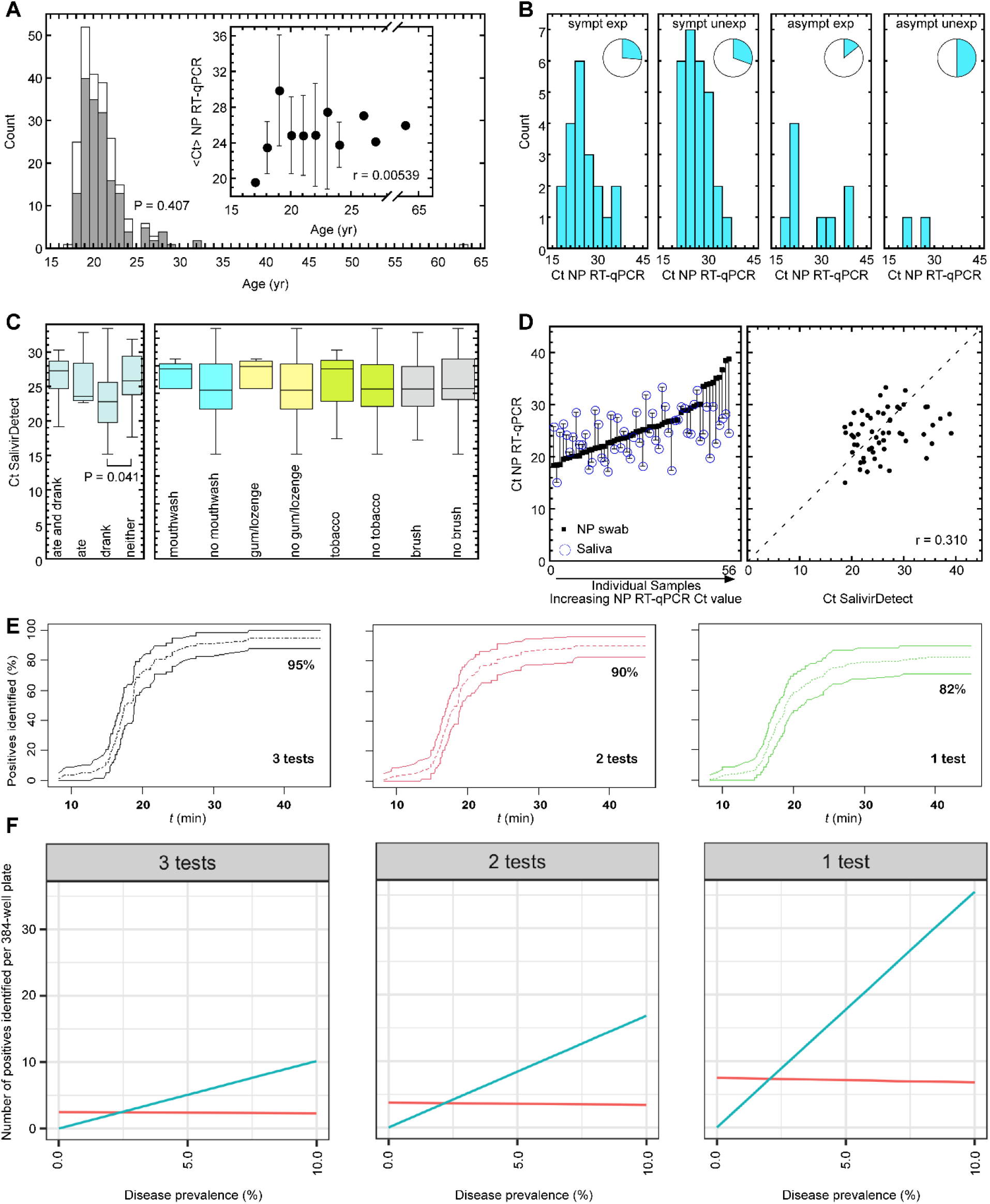
Study results and statistical predictions. Histogram of participant age distributions, N=223. Gray and white bars represent positive and negative participants, respectively, tested using CDC-approved NP RT-qPCR (p = 0.407). Inset: average Ct values of SARS-CoV-2-positive CDC-approved NP RT-qPCR tests as a function of participant age (r = 0.00539). Error bars represent one standard deviation (Panel A). Histograms of CDC-approved NP RT-qPCR Ct values of SARS-CoV-2-positive participants self-reported as (from left-to-right) symptomatic exposed (N=76), symptomatic unexposed (N=88), asymptomatic exposed (N=64), and asymptomatic unexposed (N=4). Insets: fraction of positive tests (blue) and negative tests (white) within each category (Panel B). Box-and-whisker plots of SalivirDetect Ct values for individuals who both ate and drank (N=10), ate only (N=4), drank only (N=23), and did neither (N=21) (left panel) in addition to having used mouthwash (N=5), no mouthwash (N=52), gum/lozenge (N=6), no gum/lozenge (N=52), tobacco (N=7), no tobacco (N=51), brushing (N=31), and no brushing (N=27) (right panel) within 30 min of saliva sampling. Box-and-whisker plots define minimum, first quartile, median, third quartile, and maximum values (Panel C). Comparison between collection sites on the body. The Ct values of 56 positive CDC-approved NP RT-qPCR samples are plotted in increasing order (closed black squares). Open blue circles represent the SalivirDetect Ct values of the 56 corresponding saliva samples. Vertical lines connect NP and saliva values to guide the eye (left subplot). Cluster plot of Ct values of CDC-approved NP RT-qPCR vs. SalivirDetect. Dashed line indicates perfect correlation (r = 0.310) (right subplot) (Panel D). Bootstrap-predicted percentage of total positive individuals detected within a testing population as a function of SLAMP assay time for triplicate, duplicate or single replicate test conditions. Dashed lines indicate the mean. Solid lines indicate 95% bootstrap confidence intervals (Panel E). Total number of positive cases identified using SLAMP per 384-well plate as a function of disease prevalence for three, two, and single replicate tests. True positives (blue) and false positives (red) are generated for disease prevalence rates between 0 and 10% for single replicates, duplicates, and triplicates based on a daily capacity of six 384-well plates, reserving 10 wells for controls (Panel F).

### Self-reported clinical presentations and test results

Participants were classified into four categories defined by their questionnaire responses: symptomatic exposed, symptomatic unexposed, asymptomatic exposed, and asymptomatic unexposed. Samples in each self-reported category were confirmed negative or positive based on CDC-approved NP RT-qPCR and positive individuals were assigned a Ct value. The 2nd largest category was symptomatic exposed numbering 76 individuals. Among these, only 20 participants were confirmed SARS-CoV-2-positive, around 26%. Symptomatic unexposed, the largest of all four categories, was comprised of 88 individuals from which 27 were confirmed positive, around 31%. The asymptomatic and exposed respondents numbered 64, of which only 9 tested positive, a total of only 14%. The smallest category was comprised of four participants who were asymptomatic and unexposed. Of these, half tested positive. Histograms of Ct values among the positive results with inset pie charts representing the fraction of confirmed SARS-CoV-2 positives (blue) among the negatives (white) for each category are shown in Figure 3B.

### Consumption of potential inhibitors – effect on SalivirDetect Ct values

Distributions of SalivirDetect Ct values among individuals who had or had not consumed each potential inhibitor are shown in Figure 3C. Among those engaging in eating and drinking, only the Ct values of those who drank were statistically distinct compared to those who did not (p = 0.041). No other items consumed resulted in statistically significant differences in the Ct distributions of those populations. The p-values for comparisons of all other sets were >0.05.

### Site comparison: Nasopharynx versus saliva

CDC-approved NP RT-qPCR and SalivirDetect provide Ct results that can be quantitatively compared between NP swabs and saliva (Figure 3D). We compare the viral titers of 56 individuals who tested positive under both methods. Viral titer has been shown to vary between these sites ^28^. Among samples with lower NP swab Ct values, assumed to correspond to a higher viral titer, saliva samples Ct values were higher, implying a lower viral titer than in the nasopharynx. However, when NP swab Ct values imply lower viral titers, Ct values in saliva were significantly lower, implying a difference in viral concentration between nasopharynx and oral mucosa (saliva) of possibly four orders of magnitude. Furthermore, there is a definite jump in values for the NP swab results from Ct=30-34, after which saliva Ct values are all lower than NP swab (Figure 3D, left). A correlation plot of NP vs. saliva Ct values reveals that the results have only a weak positive correlation (r = 0.310).

### Statistical predictions for SLAMP tests at triplicate, duplicate, or as a single replicate

For each number of test replicates, sensitivity increases as a function of SLAMP assay duration (Figure 3E). As reported here, 45-min SLAMP with 3 replicates produced a sensitivity of 95% (95% CI: [87.9%, 100%]) as compared to the CDC-approved NP RT-qPCR standard results. Bootstrapping predicted a 90% sensitivity (95% CI: [82.8%, 96.6%]) for SLAMP with two replicates, and a 82% sensitivity (95% CI: [70.7%, 89.7%]) for SLAMP with a single replicate after the full assay time of 45 min had passed. For example, half of all positives in a test population would be detected by a single SLAMP test after 19.0 min (95% CI: [17.7 min, 21.5 min]), duplicate tests after 18.4 min (95% CI: [17.4 min, 19.3 min]), and triplicate tests after 17.7 min (95% CI: [16.9 min, 18.8 min]).

Based upon an estimated testing capacity of six 384-well plates per day (see Methods), we compare three scenarios of running samples with a single replicate, duplicate, and triplicate (Figure 3F). For these scenarios, we use the sensitivity values from Figure 3E. The specificity is assumed to be a constant 98% for all scenarios and there are 10 wells in each plate reserved for controls. Given the fixed constraint on the number of tests (≈384 × 6), screening a larger number of individuals can identify more positive cases. For example, at a disease prevalence of 10%, triplicate SLAMP would identify approximately 10.0 true positives (TP) at the cost of 2.4 false positives (FP) per 384-well plate with FPs equaling the number of TPs at a disease prevalence of 2.5%. For duplicate SLAMP, TP = 16.8, FP = 3.3, and TP = FP at a disease prevalence of 2.2%. Finally, for single test SLAMP, TP = 35.4, FP = 6.9, and TP = FP at a disease prevalence of and 2.0%.

## Safety

Testing site volunteers were equipped with powered air-purifying respirators (PAPRs) during sample collection. Samples were transported in hard-sided coolers with biohazard labels. Contact surfaces were frequently disinfected using 70% ethanol solutions. All samples should be assumed to contain infectious SARS-CoV-2 virus and must be rendered safe through viral inactivation^29^ before testing. HEPA air purifiers (Medify Air MA-25) were used in the testing laboratory. All samples were heat inactivated before storage. See Supplementary Methods and Figure S4 for validation of viral heat-inactivation procedure.

## Discussion

The demographics of this study (Table 1) reflect those of many universities throughout the US. The median age of study participants was 20 years, and 94% of participants were ages 18-24. This study was comprised of 53% females and 47% males. By comparison, the 2020 MSU student population was comprised of 48% females, 51% males, and <1% other^30^. This difference is in agreement with observations that suggest women seek Covid-19 testing, more than males^31,32^ and are more likely to take covid precautions seriously^33^. In terms of race, this study was comprised of 94.4% white and 5.6% non-white participants. By comparison, the 2020 MSU student population was comprised of 84% white and 16% non-white^30^. This difference of 10% may reflect differences in likelihood to seek Covid-19 testing^34^. The average US university in Fall 2020 was around 49% white^35^, more racially diverse than represented within our study. Nevertheless, we have no reason to doubt that SLAMP testing would be successfully employed among the student populations of other universities.

RT-LAMP (SLAMP) and RT-qPCR (SalivirDetect) Covid-19 tests performed on the saliva of 233 adults^36^ yielded 98% specificity for both and 95% and 97% sensitivity, respectively, sensitivities comparable to SalivaDirect^37^.

Surveying revealed that while 70% of pilot study participants identified as symptomatic, only 29% of these tested positive for SARS-CoV-2 (Table 2). Overall, the prevalence of Covid-19 disease was 25%, a number that coincides with incidence levels of 9-31% across the US, as estimated in the months prior to our study^38^. Only 5% were asymptomatic positives compared to the estimates of around 40% asymptomatic among the general population^39^ which is likely due to testing being reserved only for those with symptoms or recent exposures. We expect a random screening of the population would likely have produced a higher asymptomatic positive rate, given estimates ranging from 40-45% asymptomatic positive rate^40^.

While interference from food, drink, oral care products, and tobacco are a concern for saliva diagnostics^41^, we observed no significant differences in Ct distributions of SalivirDetect positive results who had or had not consumed food, oral care products, or tobacco within 30 min of saliva sampling. Interestingly, those who had consumed drink produced significantly lower Ct values (p = 0.407) than those who had not (Figure 3C). This suggests that hydrating the mouth before expressing saliva may stimulate its production, at least temporarily^42^, and contribute to the supply of greater viral genome from the oral mucosa^43^.

Viral load in saliva is consistently higher than NP swab. Strikingly, positive samples with the highest CDC NP RT-qPCR Ct values >30 are surprisingly lower than in saliva, representing a higher concentration of virus in the saliva of the oral mucosa under conditions of low titer in the nasopharynx (Figure 3D). In fact, all saliva samples had Ct ≤ 35, and the 9 individuals with the lowest NP swab virus concentrations (Ct > 35) had significantly lower Ct for the corresponding saliva sample test. SARS-CoV-2 infections have exhibited high tissue compartmentalization and data suggests viral titers may peak earlier in saliva than in NP swabs^24^.

While the SLAMP test has higher sensitivity when run in triplicate, running a single test can be advantageous. Given finite resources, driven by cost or supply availability, a single test on an individual can identify a substantially larger amount of infected, and infectious, individuals. We modeled the sensitivities and specificities of double and single tests from the triplicate SLAMP test data. Figure 3F shows the total number of positive individuals that can be identified by running a single 384-well plate in either triplicate, duplicate, or with a single test. Triplicate and duplicate have higher sensitivity, with 95% and 90% respectively, compared to that of a single test at 82%; however, running a single test allows more individuals to be tested.

It should be noted that specificity is assumed constant across the three SLAMP replicates (Figure 3F, red lines). Therefore, running more tests, with a lower specificity can also lead to more false positives that may require confirmatory testing. Recent work^10^ has demonstrated that more frequent testing with LAMP was more effective at reducing epidemic size than deploying more sensitive tests. Our analyses regarding sensitivity predict that running more samples with single replicates will lead to more false negatives, but ideally a high frequency testing strategy would catch those other cases^10^. Nonetheless, the data suggest that the SLAMP assay run in triplicate per individual could perform at the level of a clinical diagnostic, while when run as a single replicate could be used as a broad screening tool to identify unknown positives.

These results represent the intersection of self-reported demographics, disease presentation, and potential interferences. Our study size of N=233 with 58 diseased can be statistically interpreted with acceptable accuracy^10^. However, the sample number in the case of some interferences is low and limits statistical power.

SARS-CoV-2 testing that is rapid, simple, and requires no specially trained medical personnel is indispensable when large numbers of individuals must be frequently screened while remaining economical. In this study, we have compared three testing methods, NP RT-qPCR, SalivirDirect, and SLAMP. All three methods use simple heat to both inactivate and extract, which avoids costly reagents and potential supply bottlenecks. We conducted a comparison of these tests between 233 individuals from the MSU symptomatic testing site in mid-November of 2020, at the height of the largest county-wide wave of Covid-19 cases, where we detected a disease prevalence of 25%. Compared to the “gold standard” NP RT-qPCR results, SalivirDetect and SLAMP achieved 97% and 95% sensitivity and 98% and 98% specificity, respectively. We have demonstrated our SLAMP method offers a faster and simpler, while still comparatively sensitive, alternative to NP swabs and RT-qPCR methods for routine sampling, particularly of a college population.

Supported by the State of Montana through the Coronavirus Aid, Relief and Economic Security Act (CARES) Act.

Connie B. Chang and James N. Wilking are co-founders of ULTSafety, Inc. which has licensed technology from Montana State University and Harvard University relating to this publication.

We thank Nathan Tanner at New England Biolabs, Kathryn Kundrod in the Richards-Kortum group (Rice University), Paul Hergenrother (UIUC), and the Global LAMP R&D Consortium led by Christopher Mason (Weill Cornell Medicine) for helpful discussions. We thank MSU students Kyle Hain, Shawna Pratt, Emily Walter, Matthew Fisher, and Jerrica Bursik for sample collection and Donna Gollehon (MSU University Health Partners), Ryan Brickman (MSU Safety and Risk Management), and Ryan Bartlett (MSU Office of Research Compliance) for their assistance throughout the approval process and especially with sample site logistics. We thank the MSU Center for American Indian and Rural Health Equity for the use of the Health Education and Research Bus (HERB) at the sample collection site. HERB is a mobile laboratory and outreach facility supported by an Institutional Development Award (IDeA) from the National Institute of General Medical Sciences of the National Institutes of Health under grant numbers P20GM104417 and P20GM103474.

## Data Availability

Data is available by request to corresponding authors.

## Author Affiliations

From the Montana State University Department of Chemical & Biological Engineering (D.A.B., E.K.L., T.L., I.T., J.N.W., C.B.C.), Center for Biofilm Engineering (D.A.B., C.H., K.A.B., A.P., E.K.L., T.L., I.T., J.N.W., J.M., M.W.F., C.B.C.), Department of Microbiology & Immunology (M.R., S.T.W., R.K.B., M.P.T., D.E.K., M.W.F.), Department of Mathematical Sciences (A.P., A.B.H.), Center for American Indian and Rural Health Equity (CAIRHE) (A.K.A.), and Health & Human Development (J.R.C.).

