## Supplementary Appendix for "SARS-CoV-2 Detection in the Nasopharyngeal Swabs and Saliva of College Students using RT-qPCR and RT-LAMP"

### Table of Contents

Supplementary Methods……………………………………………………………………..2
3D-printed caddy accessory for efficient saliva collection………………………..2
Duplex RT-LAMP primers for SARS-CoV-2 detection and internal control……….2
SLAMP primers……………………………………………………………………….2
SalivirDetect primers……………………………………………………………..….3
NEB Luna Kit RT-qPCR Reactions……………………………………………..….3
RT-LAMP Curve fitting……………………………………………………………….4
Synthetic vRNA standards for determining LOD……………………………………4
BSL-3 SARS-CoV-2 growth and heat inactivation of infectivity……………….…..4

Supplementary Results……………………………………………………………………..4
Optimizing saliva sample heat inactivation………………………………………...4
Comparing testing methods: SLAMP, SalivirDetect, and NP RT-qPCR……….….6
Standard curves and LOD for SLAMP reactions………………………………..…..6
RT-LAMP reaction optimized using GuHCl and duplex primer sets………………6

Supplementary Figures……………………………………………………………………8
Fig. S1. SARS-CoV-2 cases in Gallatin County, MT between Apr. 2020 - May 2021…..8
Fig. S2. Saliva collection caddy……………………………………………………..…8
Fig. S3 Saliva RT-LAMP testing workflow……………………………………………..…..9
Fig. S4. 95 °C heat-inactivation durations and subsequent SARS-CoV-2 virus infectivity…………………………………………………………………………….….9
Fig. S5. 95 °C heat inactivation duration optimization of positive saliva sample dilutions…………………………………………………………………………..…10
Fig. S6. Determining limit of detection (LOD) and standard curves for SLAMP………..11
Fig. S7. Duplex NE primer set and guanidine HCl (GuHCl) improves sensitivity and speed of SLAMP……………………………………………………………………...12

Supplementary References………………………………………………………….…13

### Supplementary Methods

#### 3D-printed caddy accessory for efficient saliva collection

To facilitate drive-through saliva collection, each subject was provided a 3D-printed caddy accessory containing a medicine cup, transfer pipet, and sample tube (Fig. S1A). Accessory designs were optimized for low cost and high throughput, enabling printing of five caddies at a time (Fig. S1B). Accessories were manufactured using an SLA 3D-printer (Formlabs, Form 2 and 3) loaded with clear UV cross-linkable resin (Formlabs, RS-F2-GPCL-04), soaked in isopropanol for 20 min, and post-cured under UV light (Formlabs, Form Cure FH-CU-01) at 60 °C for 15 min. Drainage holes were included at the bottom of each caddy to drain excess resin during printing and post-processing and drain 70% ethanol during decontamination. 3D models and print files are available as supplementary material.

#### Duplex RT-LAMP primers for SARS-CoV-2 detection and internal control

All primers were based on New England Biolabs designs[1] ordered from Integrated DNA Technologies (Coralville, IA, USA), and resuspended to 100 μM in nuclease-free water (NEB B1500L). All primers were desalted except FIP and BIP sequences, which were HPLC purified. For 100 reactions, the SARS-CoV-2 primer sets (N2, E1) were prepared as a duplex primer mix in the following manner: 40 μL of FIP, 40 μL of BIP, 10 μL of LF, 10 μL of LB, 5 μL of F3, 5 μL of B3, and 30 μL nuclease-free water (250 μL total volume). A third internal control primer set targeting human beta-actin (ACTB) was also similarly prepared, except with the addition of 140 μL of nuclease-free water (250 μL total volume). The prepared working primer solutions thus consisted of 16 μM of each FIP and BIP, 4 μM of each LF and LB, and 2 μM of each F3 and B3. The primer stocks and prepared working primer solutions were stored at -20 °C until use.

#### SLAMP primers

| Primers | Sequence 5'-3' |
|---|---|
| N2_FIP | TTCCGAAGAACGCTGAAGCGGAACTGATTACAAACATTGGCC |
| N2_BIP | CGCATTGGCATGGAAGTCACAATTTGATGGCACCTGTGTA |
| N2_F3 | ACCAGGAACTAATCAGACAAG |
| N2_B3 | GACTTGATCTTTGAAATTTGGATCT |
| N2_LF | GGGGGCAAATTGTGCAATTTG |
| N2_LB | CTTCGGGAACGTGGTTGACC |
| E1_FIP | ACCACGAAAGCAAGAAAAAGAAGTTCGTTTCGGAAGAGACAG |
| E1_BIP | TTGCTAGTTACACTAGCCATCCTTAGGTTTTACAAGACTCACGT |
| E1_F3 | TGAGTACGAACTTATGTACTCAT |
| E1_B3 | TTCAGATTTTTAACACGAGAGT |
| E1_LF | CGCTATTAACTATTAACG |

| | |
|---|---|
| E1_LB | GCGCTTCGATTGTGTGCGT |
| ACTB_FIP | GAGCCACACGCAGCTCATTGTATCACCAACTGGGACGACA |
| ACTB_BIP | CTGAACCCCAAGGCCAACCGGCTGGGGTGTTGAAGGTC |
| ACTB_F3 | AGTACCCCATCGAGCACG |
| ACTB_B3 | AGCCTGGATAGCAACGTACA |
| ACTB_LF | TGTGGTGCCAGATTTTCTCCA |
| ACTB_LB | CGAGAAGATGACCCAGATCATGT |

##### SalivirDetect Primers

| Primers | Sequence 5'-3' |
|---|---|
| 2019-nCoV_N1-F | GACCCCAAAATCAGCGAAAT |
| 2019-nCoV_N1-R | TCTGGTTACTGCCAGTTGAATCTG |
| 2019-nCoV_N1-P | **FAM**-ACCCCGCATTACGTTTGGTGGACC-**IBFQ** |
| RP-F | AGATTTGGACCTGCGAGCG |
| RP-R | GAGCGGCTGTCTCCACAAGT |
| RP-P | **Cy5**-TTCTGACCTGAAGGCTCTGCGCG-**IBRQ** |

##### NEB Luna Kit RT-qPCR Reactions

All RT-qPCR reactions used for SLAMP standard curve comparisons were prepared according to manufacturer protocol (NEB E3006L): 10 µL of Luna Universal Probe One-Step Reaction Mix (2×), 1 µL of Luna Warm Start RT Enzyme Mix (200×), 4 µL of primer mix, and 5 µL of template for a final reaction volume of 20 µL. The CDC N1 forward primer was GACCCCAAAATCAGCGAAAT, CDC N1 reverse primer TCTGGTTACTGCCAGTTGAATCTG, and probe FAM-ACCCCGCATTACGTTTGGTGGACC-BHQ1. The primer mix was prepared from a 20 µM stock of each primer and constituted the following: 0.4 µL of both forward and reverse primers (0.4 µM final concentration), 0.2 µL of probe (0.2 µM final concentration), 0.5 µL of UDG, and 2.5 µL of nuclease-free water. The reactions were performed in a QuantStudio 7 Real-Time PCR system with the following thermocycler conditions: reverse transcription at 55 °C for 10 min, initial denaturation at 95 °C for 1 min, followed by 44 cycles of denaturation at 95 °C for 10 s, extension at 55 °C for 30 s, and plate read. The data was analyzed using Design & Analysis Software, Release Version: 2.5.0, Copyright 2020 Thermo Fisher Scientific.

##### RT-LAMP Curve Fitting

RT-LAMP amplification curves were described by equation $\mathrm{Rn}(t) = a + (k - a)(1 + \exp[-b(t - m)])$ where $\Delta\mathrm{Rn}(t)$ represents a scaled and baselined fluorescence intensity as a function of reaction time $t$.[2] Fit parameters $a$ and $k$ define the lower and upper saturation limits, respectively, while $m$

and *b* describe the *x*-coordinate and slope at the inflection point, respectively. The time-to-positive Tp = *m* - 2/*b* is derived from *m* and *b* fitting parameters and serves as an analog to the familiar cycle threshold Ct used in qPCR results interpretation. Fitting was performed using KaleidaGraph v4.1 non-linear least squares routine.

##### Synthetic vRNA standards for determining LOD

Standards were prepared from heat- and TCEP-inactivated saliva to protect RNA from endonucleases and represent ideal backgrounds. Final TCEP concentrations in reactions were 2.5 mM. Full-length synthetic genomic vRNA was purchased from Twist Biosciences (Control 9 Genbank ID: MT152824, GISAID ID: USA/WA2/2020).

##### BSL-3 SARS-CoV-2 growth and heat inactivation of infectivity

Plaque assays (sensitivity limit 5 plaque-forming units (PFU)/mL) were performed on infectious solutions of USA/WA1/2020 strain (BEI Resources) SARS-CoV-2 serially diluted 10-fold in viral media which consists of 1× Dulbecco's Modified Eagle's Medium (DMEM), 2% fetal bovine serum (FBS), and 1× Pen Strep. Dilutions are inoculated onto susceptible E6-Vero cells (ATCC) and incubated for 1 h to facilitate infection. Subsequently, cells are overlaid with a diffusion-limiting agent composed of 0.75% methylcellulose and incubated at 37 °C for 3-4 days. Viral replication causes "plaques" to form on the cell monolayer, the number of which correlate with infectivity present in the original sample. Heat treatment was conducted in 1.5 mL screw cap tubes from 1.5 mL of viral supernatant at $1.2 \times 10^7$ PFU/mL and temperatures were confirmed with thermometers and digital probes[3].

### Supplementary Results

##### Optimizing saliva sample heat inactivation

Collected saliva samples may contain infectious SARS-CoV-2 virus and must be rendered safe through viral inactivation[4]. Frequently, a chemical inactivation agent is added to the sample or is present in vials prior to sample collection[5]. However, components of such inactivation buffers are often costly (tris(2-carboxyethyl)phosphine (TCEP) or thermolabile proteinase K) and have seen limited available during the COVID-19 pandemic. Chemical inactivation methods often function in a dual capacity to decrease viral infectivity[6] and to inactivate the abundant endonucleases that could rapidly degrade SARS-CoV-2 viral RNA (vRNA) and potentially inhibit downstream amplification during the RT-LAMP reaction[7]. The SLAMP method does not require the addition of any chemical agents, using instead a simple heat inactivation procedure that accomplishes viral inactivation. A range of effective temperatures and heating times have been reported[8] for sputum and other viruses[9] sometimes with chemical additives prior or post heating[10,11]. In addition, previous work has suggested that SARS-CoV-2 vRNA is stable in human saliva and detectable with molecular-based approaches[12]. However, most previous LAMP studies have used colorimetric detection with or without RNA extraction[10,13,14].

We explored heating times at a temperature of 95 °C, reported[15] to most completely inactivate SARS-CoV-2 in viral transport media. A high titer stock of SARS-CoV-2 was diluted in viral growth media in triplicate to a final volume of 200 µL. The diluted viral supernatant was then heated to 95 °C for variable lengths of time, ranging from 0 - 10 min. Infectivity was then measured via plaque assay in which a limiting dilution of virus was inoculated onto susceptible cells to determine the quantity of infectious virus present in each sample (Fig. S4A). Non-inactivated virus (Fig. S4A, solid black circles) contained $1.21 \times 10^7$ PFU/mL. The same diluted viral supernatant heated at 95 °C for 3 min (Fig. S4A, solid black squares) resulted in $3.41 \times 10^2$ PFU/mL representing a decrease of ≈$10^5$ PFU/mL. Heating for a total of 5 min resulted in a further decrease to $5.0 \times 10^1$ PFU/mL (Fig. S4A, solid black upright triangles). The final times of 7 and 10 min resulted in complete inactivation of viral infectivity with no detectable plaques at any dilution.

Similar time points were performed for infectious SARS-CoV-2 spiked into saliva that had tested negative for SARS-CoV-2 (Fig. S4B). Spiked dilutions reduced the initial infectious viral titer from $1.09 \times 10^7$ PFU/mL to $8.40 \times 10^5$ PFU/mL (Fig. S4B, solid black circles). Due to the high number of infectious virus particles present after 3 min of heating at 95 °C from the previous inactivation studies in viral supernatant, we only evaluated time points ≥5 min for saliva spiked samples. Infectious virus was undetected following exposure to 95 °C heat for 5, 7, and 10 min, indicating high titers of SARS-CoV-2 can be completely heat-inactivated in saliva in <5 min (Fig. S4B). Comparable heat inactivation of SARS-CoV-2 is only achieved at 65 °C after 20 min[3]. With the loss of some viral infectivity following the spike into saliva, we suggest a minimum heating time of 10 min at 95 °C to ensure samples are fully inactivated before use. In addition, heating at 95 °C represents a potential 2× faster option for viral inactivation compared to inactivation times reported for 65 °C.

While Infectivity experiments suggest that 5 min at 95 °C of heat inactivation is sufficient to eliminate risk of infection, the viral envelope must also be lysed to access vRNA if the most sensitive detection limits are to be achieved. Furthermore, nucleases and inhibitors present in human saliva must also be inactivated to protect free vRNA from degradation[16] while avoiding vRNA degradation due to prolonged heating. To optimize reaction conditions, we performed SLAMP reactions on raw positive saliva samples (Fig. S5). Three concentrations of SARS-CoV-2-positive saliva were prepared by dilution with negative saliva. The samples were left untreated or were heated at 95 °C for 5, 10, 15, 20, and 30 min, then amplified using SLAMP (Fig. S5A-S5C). Undiluted positive saliva (Fig. S5A) amplified under all heating durations, but relative fluorescent detection was low, $\Delta Rn < 15$. Interestingly, undiluted positive saliva added to a SLAMP reaction, heat inactivated, and lysed for the 45 min duration at 65 °C exhibited a linear, instead of sigmoidal, increase in relative fluorescence intensity ΔRn (Fig. S5A, open blue circles). At 10-fold dilution, all incubation durations resulted in sigmoidal amplification curves (Fig. S5B). The unheated positive saliva amplified the slowest (Fig. S5B, open blue circles) suggesting that the heat inactivation step is resulting in viral lysis. At 100-fold dilution, a single sample triplicate incubated at 95 °C for 5 min failed to amplify (Fig. S5C, open red squares). The mean time-to-positive Tp values <Tp> for each triplicate trial under each dilution and duration were plotted as a function of 95 °C heating time $t$ (min) (Fig. S5D). As the positive saliva sample is diluted, greater scatter is observed between amplification curves as evidenced by greater standard deviations at all heating durations (Fig. S5D, solid green diamonds). Greater scatter between triplicates is observed as the SLAMP limit of detection (LOD) is approached. 10-fold diluted samples exhibited significant differences between <Tp> without heat inactivation and at heating times $t > 5$ min (Fig. S5D, solid red squares). The undiluted saliva samples exhibit diminishing <Tp> as a function of heating time, implying detectable vRNA increased consistently from 5 – 30 min (Fig. S5D, solid blue circles). Interestingly, across all dilutions, the 30 min heating time produced <Tp> values with low standard deviation and consistent separation of ≈2 min. Similar results are observed at 20 min heating. Therefore, we choose a 15-min heating time at 95 °C for SLAMP with an additional ≈5 min to allow temperature to ramp-up, closely approximating a 20 min total heating time.

#### Comparing testing methods: SLAMP, SalivirDetect, and NP RT-qPCR

We compare samples across three different testing methods, SLAMP, SalivirDetect, and the "gold standard" RT-qPCR. Sample collection was performed using nasopharyngeal (NP) swab for RT-qPCR while saliva, shown to provide sufficient viral RNA for detection[8], was used for both SLAMP and SalivirDetect. Both SalivirDetect and NP RT-qPCR are PCR-based while SLAMP uses isothermal LAMP amplification.

#### Standard curves and LOD for SLAMP reactions

Effective tests for curbing the potential for asymptomatic or presymptomatic spread of SARS-CoV-2 must be able to detect the virus reliably and early in the course of infection before the onset of symptoms[17]. In practice, this requires a test with high sensitivity and specificity at a low limit of detection (LOD). The LOD for the "gold standard" NP qPCR method has been reported[18] as 10 copies/uL whereas publications have reported[7] the LOD and sensitivity levels for RT-LAMP assays at 2-700 copies/reaction. Furthermore, the sample matrix such as saliva, NP swab, or mid-turbinate swabs can influence the LOD due to the presence of different inhibitors, the variability in sampling methods, differences in infection between sites, or the time of sampling[19,20].

We evaluated the LOD of SLAMP using spiked saliva samples. To accurately simulate a heat-inactivated SARS-CoV-2-positive sample, we harvested saliva from an individual confirmed negative for the presence of SARS-CoV-2 using RT-qPCR and heated it for 15 min at 95 °C to inactivate endonucleases before adding full-length synthetic SARS-CoV-2 RNA (Twist Biosciences). To completely eliminate heat-resistant endonucleases, we added TCEP to saliva for a final reaction concentration of 2.5 mM. If the saliva background is not completely inactivated, synthetic RNA quickly becomes degraded and undetectable in minutes by the action of RNases[21,22] (data not shown).

Synthetic RNA-spiked samples were prepared at concentrations of $10^5$, $10^4$, $10^3$, $10^2$, 10, and 1 copies/µL and amplified using SLAMP (Fig. S6A). Amplification curves exhibit characteristic sigmoidal profiles seen in LAMP reactions, with fluorescence detection onsets ranging between 10-20 min. Each concentration is separated by ≈2 min (Fig. S6A, curves). Calculated time-to-positive Tp values are used to generate a SLAMP standard curve (Fig. S6A, inset). The apparent LOD = 1-10 copies/µL. In saliva without TCEP and only heat inactivated, the LOD is between 10-100x higher.

We provide further confirmation of contrived sample RNA concentrations by performing RT-qPCR amplification (Fig. S6B). The RT-qPCR LOD appears >1 copies/µL. The RT-qPCR standard curve (Fig. S6B, inset) spans a greater dynamic range of Ct values compared to the Tp values of RT-LAMP reactions with a significantly lower standard deviation.

#### RT-LAMP reaction optimized using GuHCl and duplex primer sets

RT-LAMP reactions involve a minimum of four primers per set to function. The addition of a loop primer pair for a total of six primers has been shown to increase the speed of the reaction. Furthermore, by including multiple 6-primer sets in a single reaction, the sensitivity and speed of the RT-LAMP reaction can be increased significantly.

Saliva is a challenging sample medium for nucleic acid testing, largely due to variability in pH[23], ionic strength , and the presence of nucleases that degrade vRNA before it can be detected[24]. RT-LAMP is more resistant to "dirty" samples, including saliva, than traditional RT-qPCR[12]. Here, we tested established methods to increase the sensitivity and speed of the RT-LAMP reaction on saliva using guanidine hydrochloride (GuHCl) and multiplexed primer sets.

Strategies to overcome sensitivity loss associated with saliva sampling include the addition of GuHCl to the RT-LAMP master mix, a strong chaotropic agent used frequently in molecular biology for varied applications including RNA isolation and protein extraction. Low GuHCl concentrations (40 mM) can be used as additives in the RT-LAMP reaction, facilitating enzyme/nucleic acid interactions and thereby increasing sensitivity without an accompanying reduction in specificity[1].

We compared triplicate SLAMP reactions of SARS-CoV-2-positive saliva using a single E1 or N2 primer set (Materials and Methods) and duplexed NE primer sets (Fig. S7A – S7D) at $10^3$ (Fig. S7A, open circles), $10^2$ (Fig. S7B, open squares), and 10 (Fig. S7C, open diamonds) genome copies/μL concentrations *C*. At $10^3$ c/μL, every primer set triplicate SLAMP reaction results in amplification and positive detection of SARS-CoV-2. At a ten-fold dilution, the E1 primer set alone begins to perform poorly, failing to amplify 2 triplicates (Fig. S7B, open blue squares). Additionally, the N2 primer set triplicates also begin to lose sensitivity at 100 c/μL. While no N2 primer set reactions failed to amplify, one reaction amplified much later (Fig. S7B, open red squares). All duplex NE primer reactions amplified (Fig. S7B, open orange squares). As the limit of detection is reached at 10 c/μL, each primer set condition drops at least a single replicate (Fig. S7C). However, the NE primer set most accurately amplifies at <Tp> appropriate for 10 c/μL (Fig. S7C, open orange diamonds). Comparison of <Tp> values for triplicates under different genome concentrations and primer sets reveal that the duplex NE primer set produces both an accurate prediction of the genome copies/uL and less variability between successful amplifications (Fig. S7D, solid green diamonds). The E1 and N2 single primer sets (Fig. S7D, solid blue circles and solid red squares, respectively) both exhibit significantly greater scatter and deviation from predicted concentrations based on the SLAMP standard curve.

After establishing that duplex NE primer sets offer greater sensitivity and reproducibility than single E1 or N2 primer sets, we evaluated whether the addition of GuHCl improved amplification reliability. We performed triplicate SLAMP reactions at positive saliva concentrations of 1000, 100, and 10 copies/uL SARS-CoV-2 genomes using the duplex NE primer sets and 40 mM GuHCl (Fig. S7E, open orange circles, squares, and diamonds, respectively). While the addition of GuHCl has partially deviated <Tp> from the standard curve, every replicate amplified and standard deviations were low (Fig. S7F).

We demonstrate that the addition of GuHCl, along with the use of a duplex primer set composed of both N2 and E1 primers (see Materials and Methods) in the same reaction, optimizes the saliva RT-LAMP reaction, achieving greater sensitivity and more rapid time-to-result, thereby increasing the utility of SLAMP for more consistent detection of low viral titers in saliva.

With an optimized assay (GuHCl, primers), knowledge of LOD and heating times, we performed our trial study. Volunteers operating at MSU's symptomatic testing site collected samples from 243 individuals who provided NP swabs for CDC RT-qPCR "gold standard" testing and saliva samples for SalivirDetect and SLAMP. A questionnaire (see Materials and Methods) was also provided to collect self-reported demographic and illness presentation data, in addition to determining whether any foods, drinks, or potential inhibitors were consumed 30 min prior to sample collection.

### Supplementary Figures

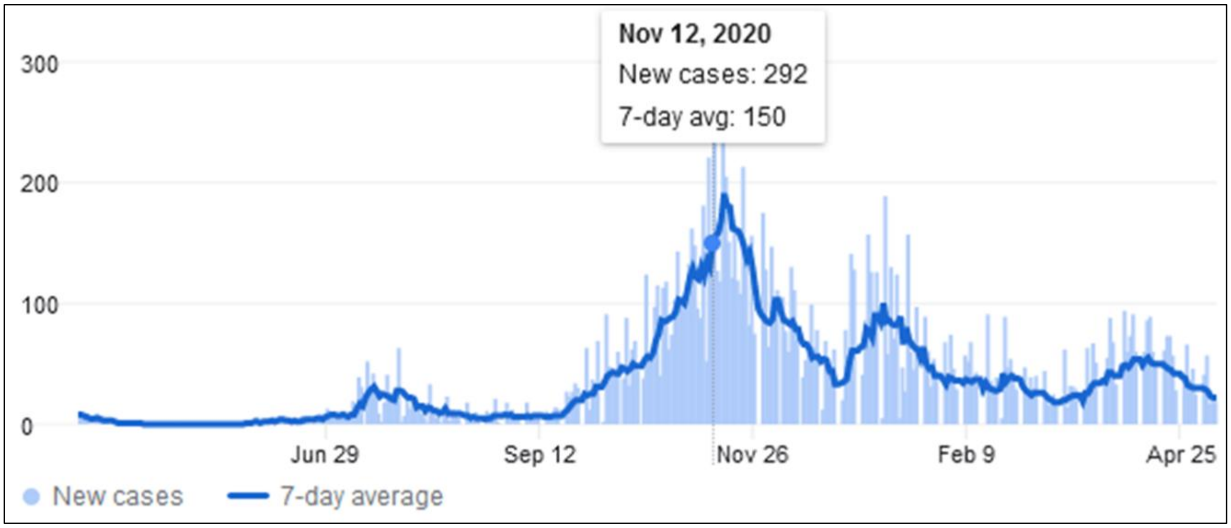


**Fig. S1. SARS-CoV-2 cases in Gallatin County, MT between Apr. 2020 – May 2021.** This comparative study was conducted at a time of peak cases (cursor and dotted vertical line). Source: The New York Times.

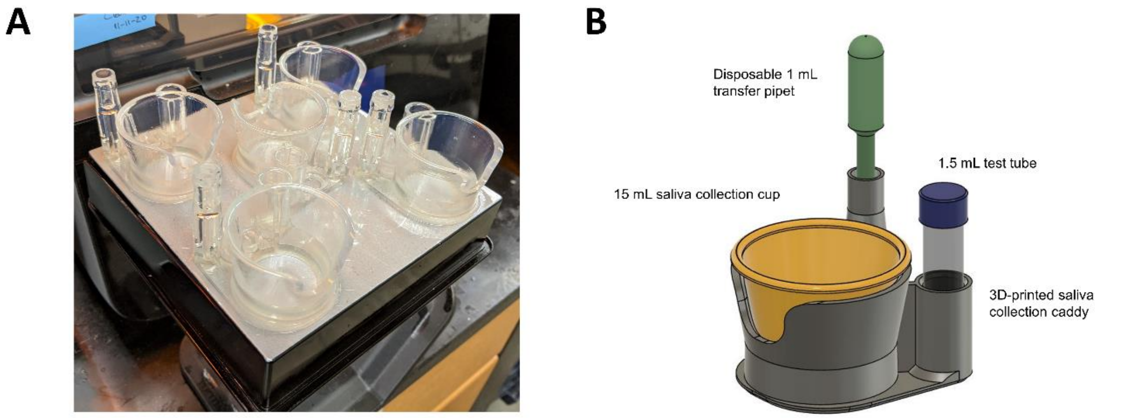


**Fig. S2. Saliva collection caddy.** (A) photograph of 5 caddies printed simultaneously. (B) A caddy as presented to a study participant, ready to collect saliva.

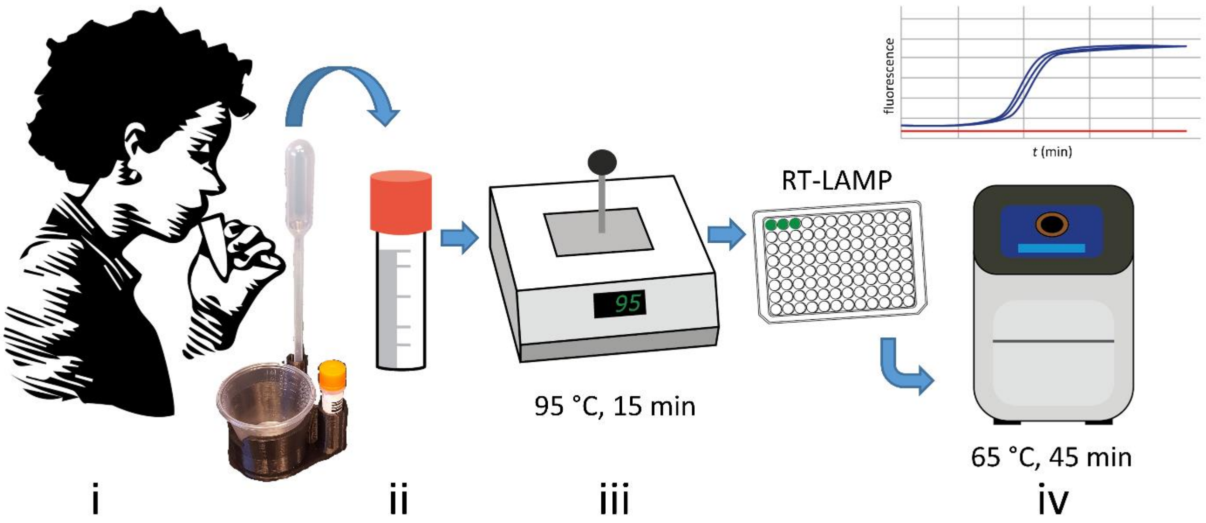


**Fig. S3. Saliva RT-LAMP testing workflow.** (i) 1mL saliva is expressed directly into medicine cup before being pipetted (ii) into screw cap sample tube. (iii) Virus inactivation is performed directly on the saliva in the tube without any added buffers in a heat block at 95 °C for 15 min. Each sample is pipetted in triplicate RT-LAMP reactions in a qPCR machine (iv) at 65 °C for 45 min.

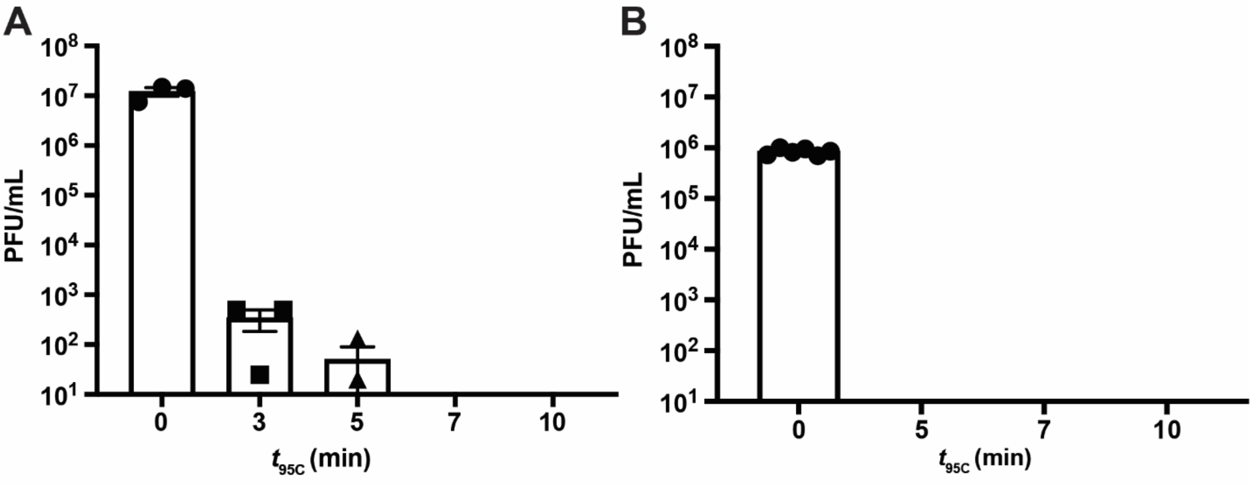


**Fig. S4. 95 °C heat-inactivation durations and subsequent SARS-CoV-2 virus infectivity.** Plaque-forming units (PFU) per mL after 95 °C heat inactivation of infectious SARS-CoV-2 for no heating (solid black circles) and 3 (solid black squares), 5 (solid black upright triangles) min in (A) viral growth media and (B) human saliva. All data represent the mean of triplicate samples +/- SEM.

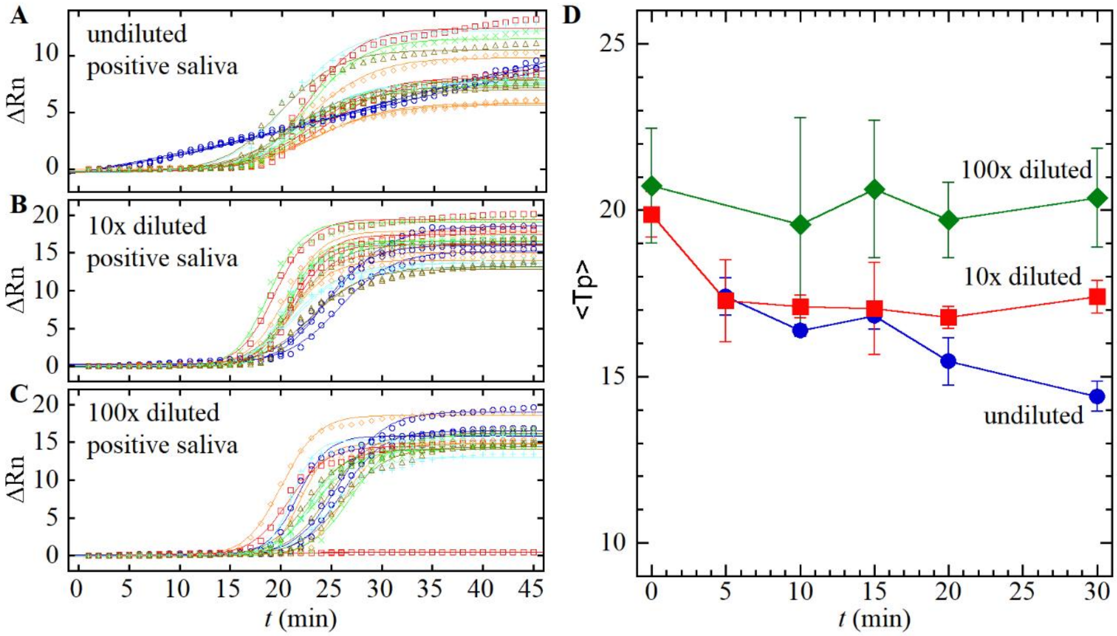


**Fig. S5. 95 °C heat inactivation duration optimization of positive saliva sample dilutions.** Triplicate SLAMP reactions were performed on 95 °C heat inactivation for 5 (open red squares), 10 (open orange diamonds), 15 (green diagonal crosses), 20 (light blue upright crosses), and 30 (open brown upright triangles) min in addition to no heating (open blue circles) for **(A)** undiluted, **(B)** 10× dilution, and **(C)** 100× dilution. Solid lines represent RT-LAMP curve fits (see Materials and Methods). **(D)** Mean time-to-positive <Tp> values as a function of 95 °C inactivation durations *t* (min) for undiluted (solid blue circles), 10× diluted (solid red squares), 100× diluted (solid green diamonds) from triplicate data in parts (A)-(C). Error bars represent a single standard deviation. Lines guide the eye.

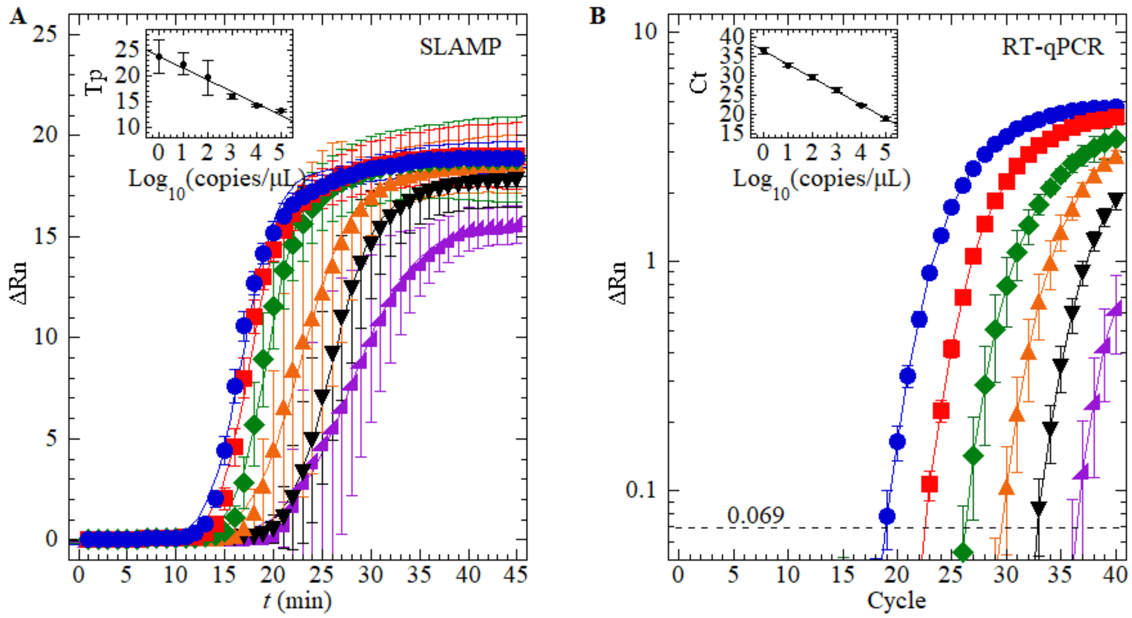


**Fig. S6. Determining limit of detection (LOD) and standard curves for SLAMP.** Contrived saliva samples prepared at $10^5$ (solid blue circles), $10^4$ (solid red squares), $10^3$ (solid green diamonds), $10^2$ (solid orange upright triangles), 10 (solid black inverted triangles), and 1 (solid purple right triangles) copies/uL. Synthetic full-length SARS-CoV-2 RNA diluted in inactivated negative human saliva and amplified using **(A)** SLAMP and **(B)** NEB Luna Kit RT-qPCR. (A) Relative fluorescence intensity ΔRn recorded as a function of time *t* (min). Limit of detection (LOD) lies between 1 and 10 c/μL. Inset: SLAMP standard curve Tp = 23.99 – 2.30$\log_{10}$ [copies/μL] ($R^2$ > 0.97) calculated using time-to-positive Tp values (see Materials and Methods) generated from data in part (A). (B) RT-qPCR standard curve. Ct values determined at ΔRn = 0.069 cutoff determined using Design & Analysis Software (see Materials and Methods). Inset: NEB Luna Kit RT-qPCR standard curve Ct = 36.55 – 3.50$\log_{10}$[copies/μL] ($R^2$ > 0.999) calculated using Ct values from part (B).

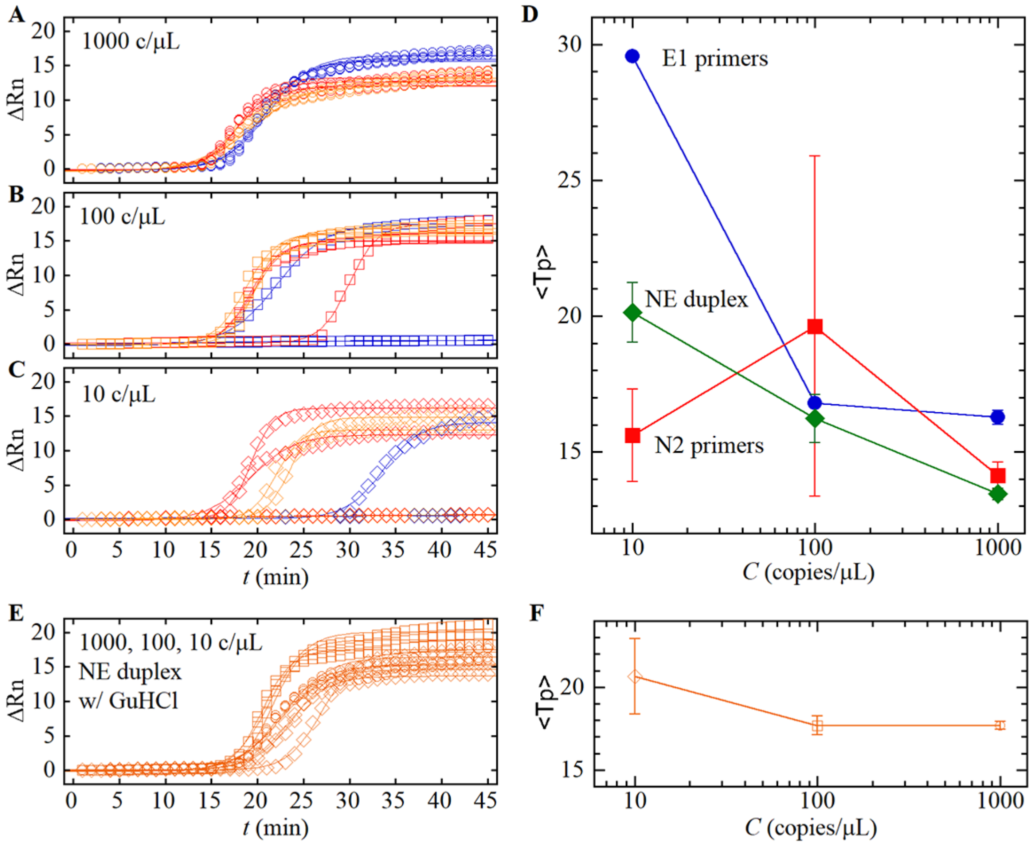


**Fig. S7. Duplex NE primer set and guanidine HCl (GuHCl) improves sensitivity and speed of SLAMP.** Triplicate SLAMP reactions of **(A)** 1000 (open circles), **(B)** 100 (open squares), and **(C)** 10 (open diamonds) copies/μL positive saliva samples without GuHCl and only single E1 primer set (blue), single N2 primer set (red), or duplex NE primer sets (orange). Curve fits yield time-to-positive Tp values (see Materials and Methods). **(D)** <Tp> as a function of genome copies/μL *C* for single E1 primer set (solid blue circles), single N2 primer set (solid red squares), and duplex NE primer sets (solid green diamonds) from parts (A) – (C). Error bars represent one standard deviation. Lines guide the eye. **(E)** Triplicate SLAMP reactions of positive saliva samples at 1000 (open circles), 100 (open squares), and 10 (open diamonds) genome copies/μL using only duplex NE primer sets (orange) with 40 mM GuHCl. **(F)** <Tp> as a function of 1000 (open circle), 100 (open square), and 10 (open diamond) genome copies/μL *C* for duplex NE primer sets with 40 mM GuHCl from part (E). Error bars represent one standard deviation. Lines guide the eye.
